# Examining the experiences of Indigenous families seeking health information for their sick or injured child: a scoping review protocol

**DOI:** 10.1101/2022.04.18.22273999

**Authors:** Lisa Knisley, Janice Linton, S. Michelle Driedger, Lisa Hartling, Shannon D. Scott

**Affiliations:** Children’s Hospital Research Institute of Manitoba, Winnipeg, Canada; Faculty of Nursing, University of Alberta, Edmonton, Canada; University of Manitoba, Neil John Maclean Health Sciences Library, Winnipeg, Canada; University of Manitoba, Rady Faculty of Health Science, Department of Community Health Sciences, Winnipeg, Canada; Alberta Research Centre for Health Evidence, Department of Pediatrics, University of Alberta, Edmonton, Alberta, Canada

**Keywords:** pediatrics, knowledge synthesis, emergency, Indigenous

## Abstract

**Introduction:** The Truth and Reconciliation Commission drew attention to the inequalities and systemic harms experienced by Indigenous peoples in Canada and called on the Canadian government and healthcare professionals to close the gap related to Indigenous communities’ access to appropriate healthcare services. The Manitoba Métis Federation (self-governing organization representing Red River Métis) identified a need for Red River Métis families to have meaningful resources when seeking emergency care for their children. A better understanding of Métis families’ experiences in seeking child health information is needed to develop culturally relevant pediatric resources. To date, the literature on Indigenous families’ experiences seeking child health information has not been synthesized. A scoping review will map the literature on Indigenous families’ experiences seeking health information to care for a sick or injured child; and identify the barriers and facilitators to accessing this information.

**Methods and analysis:** Joanna Briggs Institute methodology was used to develop the research question, *What is the extent and nature of the literature available on the experiences of Indigenous families seeking health information for their sick or injured child?* The search strategy, developed with a research librarian with extensive experience in Indigenous Peoples’ health, includes searching MEDLINE, EMBASE PsycINFO, CINAHL, and Scopus databases; grey literature, by searching the internet and consulting reference lists of key publications; examining key Indigenous research journal articles not indexed in the major biomedical databases; and snowball sampling. Two independent reviewers will screen titles and abstracts against the inclusion criteria, then screen the full texts of selected citations. Data will be extracted, collated and charted to summarize the types of studies, healthcare contexts, health information accessed, how health information was accessed, barriers and facilitators to accessing information and related measures.

**Ethics and Dissemination:** A consultation exercise with a community advisory committee will review results and inform future research. Results will be integrated with findings from other project stages to inform the adaptation of a child health resource for Red River Métis families.

## Introduction

The Truth and Reconciliation Commission [1] drew attention to the inequalities and systemic harms that have been experienced by Indigenous peoples in Canada and called on both the Canadian government and healthcare professionals to close the gap related to Indigenous communities’ access to appropriate healthcare services. A fundamental step in achieving this objective is engaging Indigenous families in developing health information that: (a) adequately and respectfully engages Indigenous families and serves their information needs; and (b) includes Indigenous knowledge and practices (e.g., rich oral traditions, experiential knowledge, and cross-cultural sharing) as core and robust sources of information. [2,3]

In this paper, we describe a protocol for a scoping review of the literature examining the experiences of Indigenous families seeking health information for their sick or injured child. This project arose from a need identified by the Manitoba Métis Federation (MMF; a self-governing organization established in 1967 representing Red River Métis people of Manitoba, Canada) for Red River Métis families to have access to meaningful and appropriate resources when seeking emergency care for their children. The Canadian Constitution recognizes Métis, First Nations, and Inuit peoples as the three Indigenous groups and first peoples of Canada. [4] A better understanding of Métis families’ experiences in seeking child health information is needed to develop culturally relevant pediatric resources. To date, the literature on Indigenous families’ experiences seeking health information for their sick or injured child has not been synthesized. The purpose of this scoping review is to map the peer-reviewed and grey literature to assess the scope of the available literature on Indigenous families’ experiences seeking child health information. Review findings will inform the cultural adaptation [5,6] of an existing child health resource to meet the identified experiences, needs and preferences of Red River Métis parents looking for health information when their child is acutely ill. Gaining a better understanding of the experiences Red River Métis parents face with regard to this decision-making process can inform how to provide accessible, useful and meaningful information to guide care. Furthermore, understanding the barriers that prevent parents from getting the information they need can help to expose and challenge existing colonial processes that are perpetuating inequitable care.

A scoping review is ideal for this purpose because it allows the incorporation of various study designs in both published and grey literature, addresses questions beyond those related to intervention effectiveness, and can be used to map the key concepts underpinning a research area. [7,8]

### Study objectives

The objectives of this review are: 1) to map the literature on Indigenous families’ experiences seeking health information to care for a sick or injured child aged 0-21 years of age, to include the transitional bridge between childhood and adulthood pediatric care [9]; and 2) to identify the barriers and facilitators to accessing this information. The research question that will guide this scoping review is: *What is the extent and nature of the literature available on the experiences of Indigenous families seeking health information for their sick or injured child?* A preliminary search of PROSPERO, MEDLINE, the Cochrane Database of Systematic Reviews, and JBI Evidence Synthesis was conducted and no current reviews on the topic were identified. Limiting the inclusion criteria to Métis may be too restrictive for this review and would not allow the exploration of what is known in an Indigenous context or to map according to any Indigenous group (e.g., Métis, First Nations, Inuit, pan-Indigenous, urban Indigenous, Maori, Native American [US context]). As such, the scoping review will be expanded to include literature relevant to other Indigenous groups and not specifically Métis.

## Methods

The scoping review will have a rigorous approach in accordance with the Joanna Briggs Institute methodology for scoping reviews,[10] which draws on the methods outlined in the Arksey and O’Malley framework [8] and augmented by Levac, Colquhoun and O’Brien [7]. PRISMA-ScR, a reporting guideline for scoping reviews [11] has informed this protocol and will also inform reporting of the subsequent scoping review findings.

### Patient and Public Involvement

The objectives and design of this scoping review were informed by a need identified by the MMF for Red River Métis families to have access to meaningful and appropriate resources when seeking emergency care for their children. This study is one stage of a multi-stage research project aimed at culturally adapting a child health resource to meet the information needs and preferences of Red River Métis parents in Manitoba. Multiple meetings have taken place with the MMF to plan the project. Integrated knowledge translation [12] activities are woven throughout the multi-stage project, including integrating end-users of the knowledge into the adaptation processes, and explicit and ongoing guidance of a community advisory committee in the research process. A presentation and/or summary of the scoping review findings will be offered to the MMF to seek feedback on the results. Additionally, a consultation exercise will take place with a community advisory committee to contextualize the results, ensure results are relevant, inform dissemination of review findings and inform future research. It is anticipated that this group will be composed of approximately 10 individuals from Métis communities (e.g., parents, Elders) in Manitoba, the MMF, and academic/health institutions (e.g., pediatric emergency clinicians, child health researchers) to provide parent and Red River Métis perspectives as well as clinical accuracy and links to healthcare systems. The results from this scoping review will inform the adaptation of the resource as well as the research engagement strategies for subsequent project stages. Patient and public involvement will be documented and reported using The Guidance for Reporting Involvement of Patients and the Public-2 short form checklist. [13]

### Eligibility Criteria

The Johanna Briggs Institute PCC mnemonic (Population, Concept, and Context) [10] was used to develop the following inclusion criteria:

1) Participants *-* Indigenous families of children aged 0-21 years. Families are defined here as parents, relatives or guardians of a child aged 0-21 years. Articles with mixed samples where data regarding Indigenous participants cannot be isolated from the larger sample will be excluded. Canada, Australia, Aotearoa New Zealand, and the United States were specifically included in this review because Indigenous Peoples within these countries share familiar stories of colonialism and its lasting impact on the health and social inequities that persist between Indigenous and non-Indigenous populations; 2) Concept - peer-reviewed and grey literature on the experiences (e.g., participation, involvement, perception, attitude) of families seeking (or accessing) health information (e.g., verbal, online or print information, materials or resources) as part of a healthcare encounter or independent of this. The study should relate to seeking health information when their child is unwell or injured; 3) Context - peer-reviewed and grey literature (that refers to a completed study) within child health (e.g., pediatrics, children/adolescents up to age 21 years, and excluding maternal/antenatal care), and includes experiences of Indigenous families seeking health information at home, in hospital or other healthcare setting.

### Information Sources

All study designs will be included to capture the full scale of the literature available in this emerging area of research. Grey literature is defined here as publicly available, open access information that may not enter normal publication or distribution channels, such as government reports, conference proceeding/abstracts and dissertations. [14] Only literature in English will be included to keep within the study budget and time restrictions. However, it is recognized that this may affect the validity of the review’s findings. [15] No date limitation will be used. A structured inclusion/exclusion form (see Supplementary File 1) will be used. The form will be pilot tested on several studies by review team members and refined, if needed, based on this pilot testing.

### Search

The search strategy, developed with a research librarian with extensive experience in Indigenous Peoples’ health (JL), includes: searching MEDLINE, EMBASE PsycINFO, CINAHL, and Scopus databases using keywords and subject headings appropriate for each database (see Supplementary File 2). Grey literature, including online dissertations from institutional repositories, will be identified by searching the internet and consulting reference lists of key publications. Additionally, a selection of Indigenous research journals will be examined to identify key journal articles which are not indexed in the major biomedical databases. Snowball sampling (backward & forward reference searching) will be carried out for relevant reviews or other key publications.

### Selection of Sources

Following the search, all identified citations will be collated and uploaded into EndNote (2013, version X9) and duplicates removed. Two independent reviewers (LK and research assistant) will screen titles and abstracts against the inclusion criteria for the review, and classify each as include, exclude or unsure. Any disagreements that arise between the reviewers at each stage of the study selection process will be resolved through discussion with a third reviewer (SS). The full text of selected citations (i.e., those classified as include or unsure) will then be retrieved and assessed in detail against the inclusion criteria (see Supplementary File 1) by two independent reviewers (LK and research assistant). The reference lists of the studies that have been selected from full-text will be screened for additional studies. “Snowball” methods, such as pursuing references of references and electronic citation tracking (e.g., Google Scholar, Web of Science, Scopus) will also be used to identify relevant sources. [16] Authors will be contacted for clarification of information (e.g., unpublished results) as required. Reasons for exclusion of full text studies that do not meet the inclusion criteria will be documented. A PRISMA-ScR flowchart will present how many studies were identified and selected, which will be accompanied by a narrative description of the search decision process.

### Data Charting Process

Data will be extracted from relevant studies by two reviewers using a structured data extraction form developed in MS Excel. For example, citation details, publication type, study design, participant details (i.e., age, sex, location), healthcare setting and context (e.g., health condition), types of health information accessed, how health information was accessed, barriers and facilitators to accessing information and related measures will be extracted and documented (see Supplementary File 3). The form will be pilot tested on three articles to allow reviewers to ensure all relevant data are extracted [8]. The form may be modified if needed during the process of extracting data from each included study. Any disagreements that arise between the reviewers will be resolved through discussion, or with a third reviewer. Study authors will be contacted to request missing or additional data, where required.

### Critical Appraisal

Unlike systematic reviews, scoping reviews provide an overview of the existing evidence, regardless of quality. [17] A formal assessment of methodological quality/risk of bias is not congruent with the purpose of scoping reviews [17] and will not be done as part of this review.

### Synthesis of Results

The analysis of the extracted data will be descriptive, and we will look for patterns and trends. The extracted data will be presented in tabular form. Additionally, a summary of the types of study designs (e.g., qualitative, quantitative), healthcare contexts, types of health information accessed, how health information was accessed, barriers and facilitators to accessing information and related measures will be presented in a narrative format.

Best practices in reporting scoping reviews, including Joanna Briggs Institute guidelines [10] and the PRISMA-ScR Checklist [11] will be followed. Review findings will be submitted to a peer-reviewed journal and presented at conferences. Review results will be used to inform the adaptation of a child health resource, as well as future research and engagement planning.

## Data Availability

All data produced in the present study are available upon reasonable request to the authors

## Ethics Approval

Ethical approval is not applicable as this review will be conducted on published literature only.

## Authors’ contributions

*All authors contributed to study design. L.K drafted the manuscript and J.L*., *S.M.D*., *L.H*., *S.S. made substantive revisions to it*.

