## Supplementary File 2 for "Examining the experiences of Indigenous families seeking health information for their sick or injured child: a scoping review protocol"

### Medline

Database: Ovid MEDLINE(R) and Epub Ahead of Print, In-Process, In-Data-Review & Other Non-Indexed Citations and Daily <1946 to current>

Search Strategy:

--------------------------------------------------------------------------------

1 exp indigenous peoples/ or american native continental ancestry group/ or alaska natives/ or indians, north american/ or inuits/ or Oceanic Ancestry Group/

2 (Indigenous or First Nations or First Nation or Inuit* or metis or Aboriginal or Native American* or alaska native* or (Alaska* adj1 Native*) or Ojibw* or Cree or Athapaskan or Athabaskan or Athabascan or Saulteau* or Wakashan or Dene or Inuk or Tlicho or Haida or Ktunaxa or Tsimshian or Gitsxan or Nisga'a or Haisla or Heiltsuk or Oweenkeno or Kwakwaka'wakw or Nuu chah nulth or Tsilhqot'in or Dakelh or Wet'suwet'en or Sekani or Dunne-za or Dene or Tahltan or Kaska or Tagish or Tutchone or Nuxalk or Salish or Stl'atlimc or Nlaka'pamux or Okanagan or Sec wepmc or Tlingit or Anishinaabe or Anishinabe or Blackfoot or Nakoda or Tasttine or TsuuT'ina or Gwich'in or Tagish or Tutchone or Algonquin or Algonkian or Nipissing or Kahnawake or Mohawk* or Cherokee or Potawatomi or Innu or Maliseet or Mi'kmaq or Passamaquoddy or Haudenosaunee or Cayuga or Lakota or Navajo or Zuni or Hopi or Oneida or Onondaga or Seneca or Tuscarora or Wyandot or Indigeneity or Nunavut* or Iqaluit* or Nunavummiut or Kitikmeot or Kivalliq or qikiqtani or Baffin or Kuujjuaq or Inuvialuit or Nunavik or nunatsiavut or Inupiat or inupiaq or innu or yupik or Yellowknife or northwest territories or Yukon or Whitehorse or Fairbanks or quajigiartiit or eskimo* or maori* or torres strait island* or koori or goori or murri or nyoongah or Nyoongar or Noongar or Nyunga* or koorie or yolngu or Anangu or palawa or nunga or Ngarrindjeri or murray island or mer island or american Indian* or aborigine* or indigen* or Hawaii* or ha waii* or Menominee or Ahousat or Apache tribe or Arapahoe or Bella Coola or Paiute or Shoshone or Blackfeet or Cherokee or Cheyenne or Sioux or Siouan or Choctaw Indian* or Choctaw Nation* or Comanche nation or Kootenai Tribes or Dogrib or Flathead Nation or Flathead Reservation or Havasupai or Pima or Tohono O'odham).mp.

3 Health Services, Indigenous/

4 1 or 2 or 3

5 exp adolescent/ or exp child/ or exp infant/ or (infant disease* or childhood disease*).ti,ab,kf. or (adolescen* or babies or baby or boy? or child* or girl? or infant* or juvenil* or kid? or minors or minors* or neonat* or neonat* or newborn* or new-born* or paediatric* or pediatric* or pediatric* or

perinat* or preschool* or puber* or pubescen* or school* or teen* or toddler? or underage? or under-age? or youth* or offspring).ti,ab,kf.

6 (parent* or family or families or guardian* or mother* or father* or grandparent* or grandmother* or grandfather* or carer* or caregiver* or lay person* or lay people* or lay man*).ti,ab,kf.

7 consumer health information/ or health literacy/ or patient education as topic/ or teach-back communication/

8 information seeking behavior/ or literacy/ or consumer behavior/

9 exp access to information/ or exp patient rights/ or right to health/ or patient advocacy/ or help seeking behavior/

10 communication/ or exp communication barriers/ or exp computer literacy/ or exp disclosure/ or exp health communication/ or exp information dissemination/ or exp information literacy/ or internet access/ or "internet use"/ or exp social networking/

11 publications/ or pamphlets/ or information services/ or exp library services/

12 exp Communications Media/

13 exp attitude to health/ or health knowledge, attitudes, practice/ or "patient acceptance of health care"/ or patient participation/ or exp patient satisfaction/

14 exp Health Services Accessibility/

15 exp "Quality of Health Care"/ or Cultural Competency/

16 professional-patient relations/ or duty to recontact/ or nurse-patient relations/ or physician-patient relations/

17 hospital records/ or exp medical records/ or nursing records/ or disease notification/

18 (disclos* or navigat* or discharge* or website* or web site* or book or books or library or libraries or blog* or support group* or peer* or social media or app or apps or phone* or telephone* or smartphone* or iphone* or android or mobile device* or podcast* or youtube or video or videos or videorecording* or audiovisual or Online or internet or facebook or written or writing or advice or helpline* or hotline* or leaflet* or brochure* or instruction* or information or pamphlet* or booklet*).ti,ab,kf.

19 or/7-18

20 health services/ or adolescent health services/ or exp child care/ or exp community health services/ or exp emergency medical services/ or health services for persons with disabilities/ or exp mental health services/ or exp nursing care/ or exp nursing services/ or exp patient care/ or exp patient escort service/ or exp personal health services/ or exp pharmaceutical services/ or exp rehabilitation/ or exp rural health services/ or exp urban health services/

21 (hospitaliz* or hospitalis*).ti,ab,sh.

22 20 or 21

23 4 and 5 and 6 and 19 and 22

24 limit 23 to english language

25 limit 24 to yr="1860 -Current"

***************************

**CINAHL on EBSCOhost**

(MH "Indigenous Peoples+") OR (MH "Aboriginal Canadians+") OR (MH "Eskimos+") OR (MH "First Nations of Australia+") OR (MH "Maori") OR (MH "Native Americans") OR “first nation” OR “first nations” OR indigenous OR aboriginal OR metis OR inuit* OR Nunavut OR iqaluit OR Nunavik OR Yukon OR Whitehorse OR Yellowknife OR “northwest territories” OR “native American*” OR “alaska native*” OR “Alaskan native*” OR “native Alaskan*” OR maori OR “torres strait islander*” OR “american Indian*” OR aborigine* OR indigen* OR hawaii* OR “ha waii” OR Hawaiian OR (MH "Medicine, Native American") OR (MH "Medicine, Traditional") OR (MH "Health Services, Indigenous")

AND

(MH "Information Resources+") OR (MH "Medical Records+") OR (MH "Health Education+") OR (MH "Telecommunications+") OR (MH "Computers and Computerization+") OR (MH "Professional-Patient Relations+") “consumer health” OR “health literacy” OR communication OR disclos* OR navigat* OR discharge* OR “patient education” OR website* OR “web site*” OR book OR books OR library OR libraries OR blog* OR “support group*” OR peer* OR “social media” OR app OR apps OR phone* OR telephone* OR smartphone* OR iphone* OR android OR “mobile device*” OR podcast* OR youtube OR video OR videos OR videorecording* OR audiovisual OR Online OR internet OR facebook OR brochure* OR instruction* OR pamphlet* OR booklet*

AND

(MH "Parents+") OR (MH "Guardianship, Legal+")  OR  parent* OR family OR families OR guardian* OR mother* OR father* OR carer* OR caregiver* OR "lay person" OR "lay people" OR "lay man*"

AND

(MH "Adolescence+") OR (MH "Child+") OR (MH "Infant+") OR (MH "Minors (Legal)") OR pediatric* OR paediatric* OR child* OR youth* OR adolescen* OR "young adult*" OR teen* OR offspring

**Sources consulted [JL] for keywords and search terms for Indigenous People**

Appropriate Terminology, Representations, and Protocols of Acknowledgement for Aboriginal and Torres Strait Islander Peoples. [Internet]. Bedford Park, SA.: Flinders University; 2012. Available from: https://dlb.sa.edu.au/tlsmoodle/pluginfile.php/14777/mod_folder/content/0/2_%20Appropriate%20Terminology%2C%20Indigenous%20Australians.pdf?forcedownload=1

Campbell, S, Dorgan, M, Tjosvold, L. Rev. March 8, 2016. [Internet]. [Edmonton]: John W. Scott Health Sciences Library, University of Alberta. Available from: https://guides.library.ualberta.ca/health-sciences-search-filters/indigenous-peoples

King, J, Masotti, P, Dennem, J, Hadani, S, Linton, J, Lockhart, B, et al. The Culture Is Prevention Project: Adapting the Cultural Connectedness Scale for Multi-Tribal Communities. Am Indian Alsk Native Ment Health Res. 2019;26(3):104-135. doi:10.5820/aian.2603.2019.104

Lowitja Institute. LIt.Search [Internet]. Carlton South, Vic.: The Lowitja Institute; 2020. Available from: https://www.lowitja.org.au/page/research/lit-search

Native Health Database [Internet]. Albuquerque, NM: University of New Mexico, [nd]. Available from: https://hslic-nhd.health.unm.edu/
