## Supplementary File 1 for "Examining the experiences of Indigenous families seeking health information for their sick or injured child: a scoping review protocol"

**Draft Inclusion/Exclusion Screening Form**

First Author, Year:________________________________________________
Article title: _______________________________________________________
Article language (only English literature included): __________________________________
Publication type (must be research; not opinion pieces or editorials): ______________________
Study published year: ________________
**Assessor's initials: __________________Date of screening: ____________________**

| **Primary Inclusion/Exclusion Criteria** | | | **(circle one for each row)** |
|---|---|---|---|
| 1. Relevant to scoping review topic | Yes | No | Cannot Determine |
| 2. Participants include *Indigenous* families in North America, Australia or New Zealand (e.g., Metis, First Nations, Inuit, Native American, Aboriginal, Maori). *Families* include parents, relatives or guardians of a child aged 0-21 years. | Yes | No | Cannot Determine |
| 3. Relates to the *experiences* (e.g., participation, involvement, perception, attitude) of families *seeking* (or accessing) *health information* (e.g., verbal, online or print information, materials or resources) as part of a healthcare encounter or independent of this. Study should relate to seeking health information when their child is unwell or injured. | Yes | No | Cannot Determine |
| 4. Literature relates to *child health* (e.g., pediatrics, children/adolescents up to age 21 years). Exclude maternal/antenatal care (health during pregnancy or childbirth). | Yes | No | Cannot Determine |
| 5. The study relates to seeking health information at home, in hospital or other setting where health care is provided (e.g., primary care, school, dental office) | Yes | No | Cannot Determine |
| 6. Is a completed research study or refers to a completed study (e.g., grey literature - government reports, dissertation, conference proceedings/abstracts), but not an editorial, opinion article or commentary | Yes | No | Cannot Determine |

Retain for (check all that are applicable):
_____BACKGROUND/DISCUSSION with review team
_____REVIEW OF REFERENCES
_____Other_______________________________
