## Supplementary File 3 for "Examining the experiences of Indigenous families seeking health information for their sick or injured child: a scoping review protocol"

### Draft Data Extraction Form

| Scoping Review Details |
|---|
| **Study Details and Characteristics** |
| Study citation details (e.g. author(s), date, title, journal, volume, issue, pages) |
| Publication type (e.g., published research, dissertation, conference abstract) |
| Geographic location of the study (or, if not listed, the location affiliation of the first author) |
| **Methodology** |
| Study design (e.g., qualitative, quantitative, mixed methods) |
| Methods of data gathering |
| **Participants** |
| Family member description (e.g., parent, sibling, guardian, etc.) |
| Total number included in analysis |
| Age |
| Gender |
| Indigenous Group |
| Country/Region/Town/Indigenous community located |
| **Context of interest** |
| Home or healthcare setting (e.g., acute care, school, dental office, primary care, home care) |
| Child health area/discipline |
| **Details/Results extracted from study** (in relation to the concept of the scoping review) |
| Type of health information/resource assessed |
| How did participants access health information? |
| Reported barriers to accessing information |
| Reported facilitators to accessing information |
| How were barriers/facilitators measured? |
